# Prison population reductions and COVID-19: A latent profile analysis synthesizing recent evidence from the Texas state prison system

**DOI:** 10.1101/2020.09.08.20190884

**Authors:** Noel Vest, Oshea Johnson, Kathryn Nowotny, Lauren Brinkley-Rubinstein

**Author notes:** corresponding author Department of Psychiatry and Behavioral Sciences, Stanford University School of Medicine Address: 1070 Arastradero Rd. Ste 200 Palo Alto, CA 94304.

## Abstract

**Importance:** People in prison are particularly vulnerable to infectious disease due to close living conditions and the lack of protective equipment. Public health professionals and prison administrators seek information to guide best practices regarding prison population to capacity rates for the COVID-19 outbreak.

**Objective:** Using latent profile analysis, we sought to characterize Texas prisons on levels of COVID-19 cases and deaths among incarcerated residents, and COVID-19 cases among prison staff.

**Design:** This observational study was a secondary data analysis of publicly available data from the Texas Department of Criminal Justice (TBDJ). Data were downloaded and analyzed on July 24, 2020. This project was completed in collaboration with the COVID Prison Project.

**Setting:** One-hundred and three prisons in the state of Texas.

**Participants:** The unit of analysis is the individual prison units that comprise the TDCJ.

**Exposures:** None

**Main Outcomes and Measures:** Latent profiles on levels of incarcerated resident COVID-19 cases, staff COVID-19 cases, and incarcerated resident COVID-19 deaths.

**Results:** We identified relevant profiles from the data: a low outbreak profile, a high outbreak profile, and a high death profile. Additionally, current prison population and level of employee staffing predicted membership in the high outbreak and high death profiles when compared to the low outbreak profile.

**Conclusions and Relevance:** Housing persons at 85% of prison capacity may minimize the risk of infection and death related to COVID-19. Implementing this 85% standard as an absolute minimum should be prioritized at prisons across the US.

**KEY POINTS:** *Question:* Is there a population to capacity ratio for prisons to successfully reduce the number of COVID-19 infections and deaths among its incarcerated populations?

*Findings:* The prisons that were most effective in reducing prison outbreaks and deaths operated at 85% of their current capacity.

*Meaning:* Prisons should operate at 85% of capacity or less to successfully minimize the harmful effects of COVID-19 on incarcerated populations.

## INTRODUCTION

Due to their close living conditions and limited opportunity for physical distancing, people in prisons are extremely vulnerable to COVID-19 infection.^1^ As a result, prisons have become hotspots for recent COVID-19 outbreaks.^2^ There has been a 21.4 percent increase in COVID-19 cases in prisons from July 13, 2020, to July 26, 2020, such that persons incarcerated are infected at nearly 4 times the rate of the general public, and prison staff are infected at two and half times the rate of the general public^3^. Despite the recent surge, very little is known regarding the characteristics of prisons with high outbreaks or high deaths. A better understanding would provide prison administrators, researchers, and health care professionals with valuable information for public health policy and planning as it relates to COVID-19 infections in state prisons.

We use latent profile analysis (LPA) to provide data-driven patterns of the COVID-19 outbreak in Texas Department of Criminal Justice (TDCJ), the largest state prison system. TDCJ has the highest level of COVID-19 cases and deaths in the nation and reports active cases at 97% of the prison facilities.^3^ TDCJ provides a unique opportunity to examine data patterns because all residents and staff have been tested for COVID-19.^4^

## METHOD

We used publicly available data from TDCJ and COVID Prison Project^4^ collected up to July 24, 2020. The primary outcome was a latent profile of Texas prisons based on their levels of incarcerated resident COVID-19 cases, incarcerated resident COVID-19 deaths, and staff COVID-19 cases. Secondary outcomes included prison level predictors of latent profile membership (population, capacity, age of the prison, and staff levels). We excluded three prison facilities because they were identified as holding facilities for individuals that had recently violated parole. Our final sample included 103 Texas prison facilities.

### Data analysis

We analyzed the data using MPlus version 8.3.^5^ First, LPA models were evaluated to determine the profile structure. We used LPA to group for patterns in the data based on three continuous observed indicators: 1) reported COVID-19 cases among incarcerated individuals, 2) reported COVID-19 deaths among incarcerated individuals, and 3) reported COVID-19 cases among prison staff (**Figure 1**). We planned to include staff deaths, but the relatively low level of staff deaths created convergence problems.

**Figure 1.**
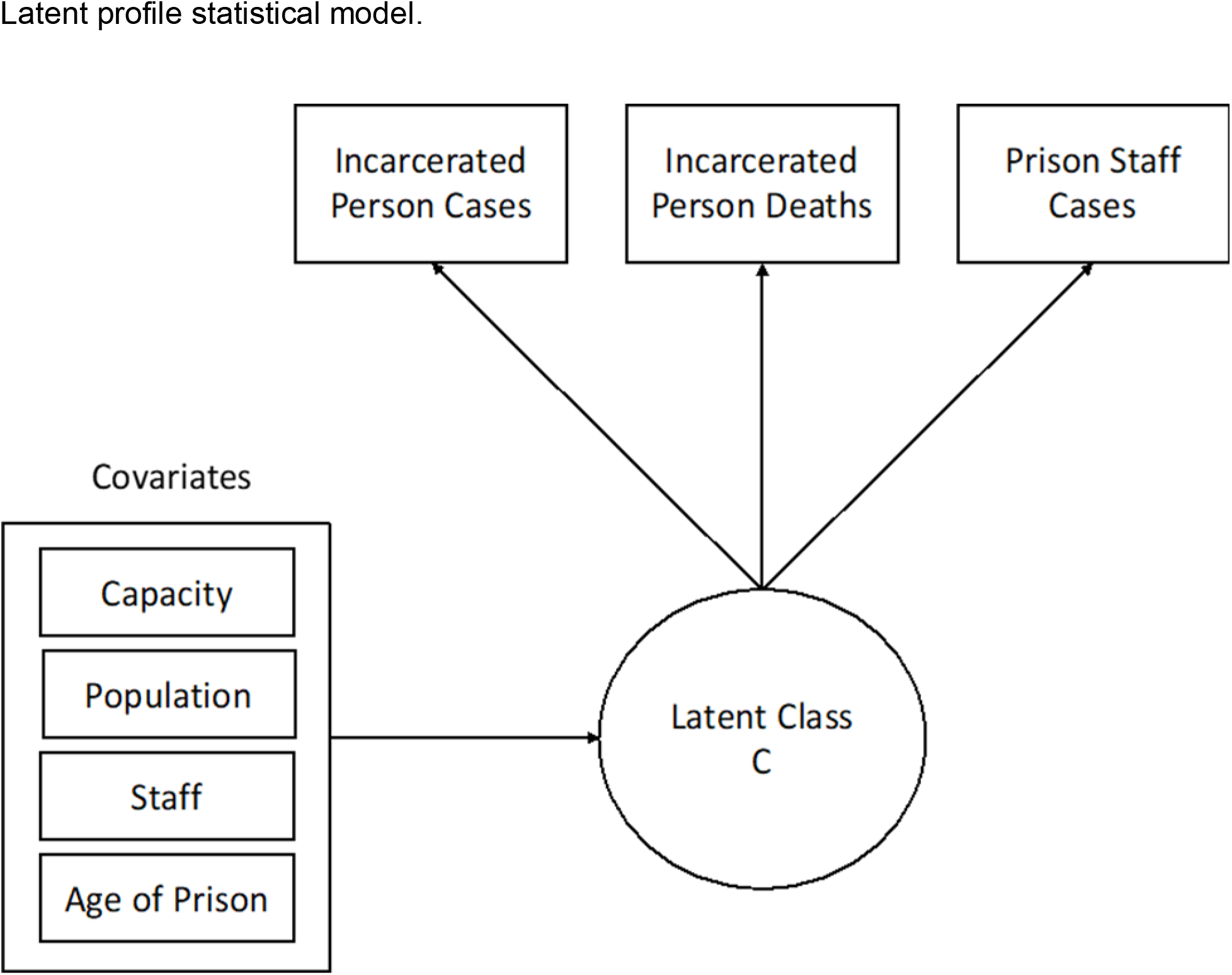
Latent profile statistical model.

We estimated 1–5 latent profile solutions. Model selection was based on standard fit statistics (BIC, entropy scores, and LRT scores).^6,7^ Once the latent profiles were identified, we examined the association between covariates and profiles using a model-based multinomial logistic regression.^8^ The three-step method was preferred because it produces more stable and less biased estimates with small sample sizes (e.g., 100–200).^9^ Each of the predictor covariates were entered into the model separately. For all of our logistic models, we used profile 1 as the referent profile. We had no missing data.

### Ethics

Because the data was publicly available, it did not require approval from the University Institutional Review Board.

## RESULTS

There were 11,799 confirmed COVID-19 cases within TDCJ among incarcerated residents, 104 presumed COVID-19 deaths among incarcerated residents, 2,497 confirmed cases among prison staff, and 12 deaths among prison staff. The entire resident population of the prisons included in the analyses was 130,610 and the entire staff population was 37,201.

We identified the three-profile solution as the most parsimonious (BIC 2595.08, entropy.99, non-significant LMR). Profile 1 (88 out of 103 prisons – dashed black line in **Figure 2**), “low outbreak” facilities, was characterized by prisons with a low number of incarcerated resident cases, a low number of incarcerated resident deaths, and a low number of prison staff cases. Profile 2 (5 out of 105 prisons – black line in **Figure 2**), “high death” facilities, was characterized by prisons with a moderate number of incarcerated resident cases, a very high level of incarcerated resident deaths, and a high level of prison staff cases. Profile 3 (10 out of 103 prisons – grey line in **Figure 2**), “high outbreak” facilities, was characterized by prisons with a very high level of incarcerated resident cases, a moderate level of incarcerated resident deaths, and a high level of staff cases. In **Supplement 1**, we offer a listing of the prisons in each profile.

**Figure 2.**
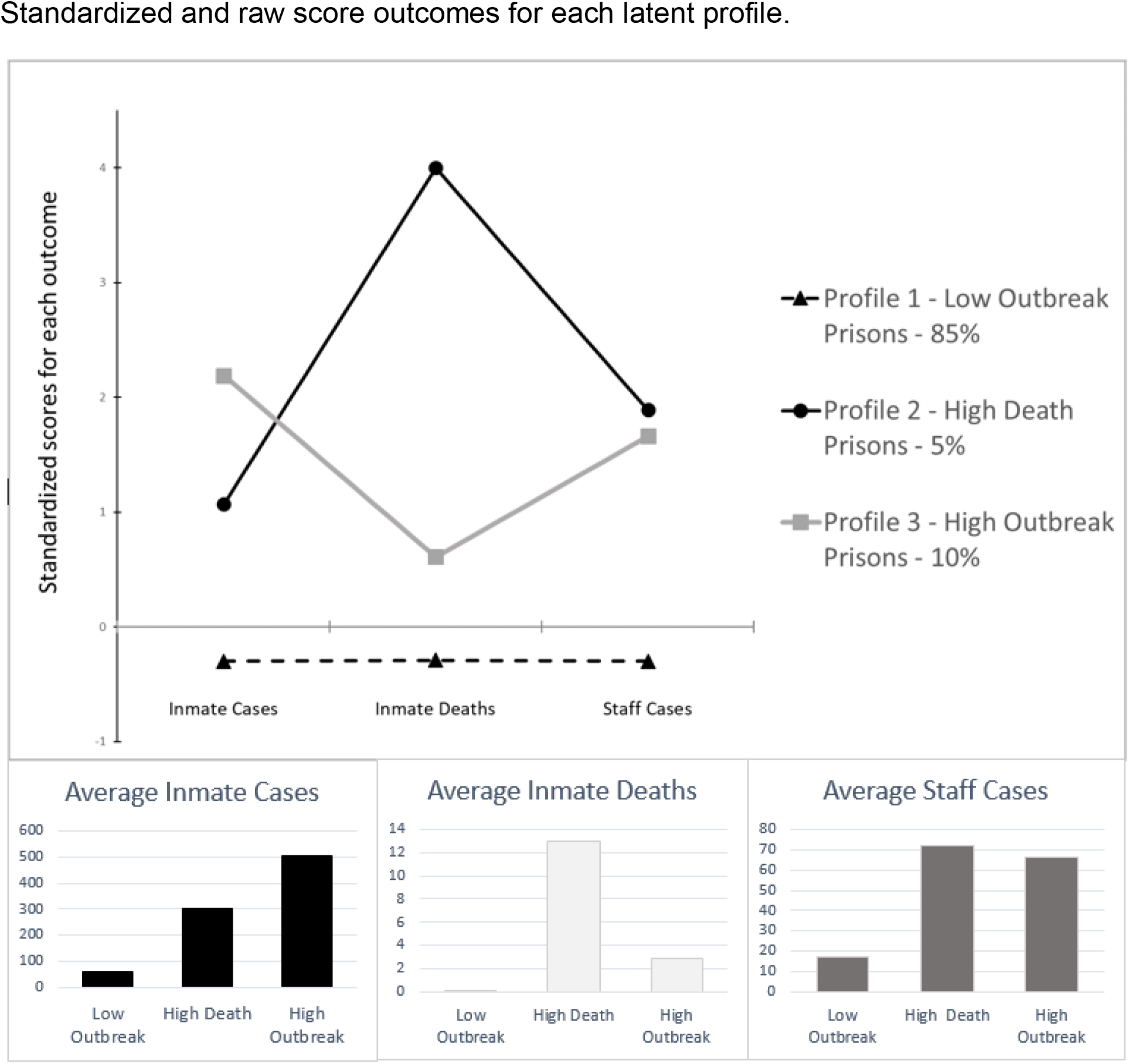
Standardized and raw score outcomes for each latent profile.

The effects of covariates on profile membership were analyzed for differences from profile 1 (low outbreak; **Table 1**). Current prison population significantly predicted membership in the high outbreak and high death profiles when compared to the low outbreak profile. Low outbreak prisons were at 85% of capacity, while the high death and high outbreak profiles were at 94% and 102% capacity, respectively. Current number of employees significantly predicted membership in the high outbreak and high death profiles compared to the low outbreak profile. We found no statistical differences among profiles in age of operation for a prison.

**Table 1.** Prison characteristic and risk covariates.

| Variable – Mean/% (SE) | Profile 1 (*referent*) | Profile 2 | Profile 3 |
| --- | --- | --- | --- |
| Current Facility Capacity | 1238.28 (77.34) | 2016.60 (490.96) | 2568.11* (341.63) |
| Current Facility Population | 1077.77 (75.65) | 1934.60* (472.73) | 2603.90* (299.52) |
| Population to Capacity Ratio <sup>a</sup> | 85% | 94% | 102% |
| Employees | 315.68 (18.08) | 571.20* (149.26) | 656.50* (73.91) |
| Years in Operation | 37.98 (3.18) | 52.60 (21.22) | 36.70 (3.44) |

Note = \* odds ratio indicates significant difference from the referent group. <sup>a</sup> denotes that this ratio was included for explanatory purposes only and was not included in the analysis of statistical differences between profiles. Profile 1 = Low Outbreak; Profile 2 = High Death; Profile 3 = High Outbreak.

## DISCUSSION

This is the first study to examine COVID-19 cases in a statewide prison system. We found that the majority of prisons in Texas were characterized by low levels of COVID-19 outbreaks among staff and incarcerated residents. Additionally, the level of overcrowding in the low outbreak prisons was moderate with a current population to capacity ratio of 85%. This suggests that the benchmark for prisons to effectively reduce COVID-19 infections should be set to under 85% of capacity. Importantly, this 85% standard should be implemented as an absolute minimum rate.

More than half of the total number of COVID-19 deaths in Texas were attributed to five prisons (65 out of 103 deaths) and nearly half of the total cases of COVID-19 were attributed to 10 prisons (5,000 out of 11,799 people). This suggests that there are COVID-19 prison hotspots, which may be connected to the overcrowding issue, understaffing, or other common characteristics that facilities share, such as resident demographics. For example, two prisons in the “high death” profile are geriatric facilities. In this inherently overcrowded environment, we suggest prisons should continue to drastically reduce their prison populations through decarceration efforts as a best practice to mitigate harms.^10^ This is especially true for those over 55 years of age^11^ since there is a growing need for gerontological knowledge and skills in prisons to help mitigate the growing infection and death rates occurring among the older incarcerated population.^12^ Lastly, age of the prison was not predictive of profile membership which suggests that building new prisons will not mitigate the public health crisis of COVID-19 infections in the prison environment.

### Limitations

This study has several limitations. First, we did not account or control for other potentially important prison characteristics. Second, the current study only examines Texas prison facilities and may not be generalizable to other state prison systems. Third, data is updated daily, therefore the number of tests, cases, and deaths of incarcerated individuals and staff will change as time progresses and our results only captures data up to July 24, 2020.

## Conclusions

We implemented a unique data-driven statistical technique to divide prisons into clusters based upon reported levels of infections and deaths at the state level. These findings should inform researchers, prison administrators, lawmakers, public health officials, and other professionals interested in reducing the impact of COVID-19 in our nation’s prisons. Importantly, housing people incarcerated at 85% of facility capacity may minimize the rate of infection and death in state prisons.

## Data Availability

These data are publically available from the COVID Prison Project and available from the authors on request.

https://covidprisonproject.com/

## Notes

### Competing Interest Statement

The authors have declared no competing interest.

### Funding Statement

Funding: Dr. Vest was supported by the National Institute on Drug Abuse of the National Institutes of Health under Award Number T32DA035165; The COVID Prison Project is funded by the Langeloth Foundation

